# Bias-domain triangulation of non-convergent observational evidence in mental health research

**DOI:** 10.64898/2026.07.07.26357342

**Authors:** Xuanyu Shi, Guanghui Deng, Jian Du

## Abstract

Observational estimates in mental health can diverge because exposure is structured by familial, clinical and social factors. We developed bias-domain triangulation to test whether this divergence follows specific sources of confounding. The framework separates reported adjustment sets from an independently generated causal structure, assigns variables to causal roles and maps back-door pathways into bias domains before pooling. We applied it to prenatal paracetamol exposure and offspring autism spectrum disorder or attention-deficit/hyperactivity disorder. The review included 24 articles and 39 adjusted estimates. The pooled association declined from 2.08 under weak overall control to 0.98 under strong overall control. Only strong familial/genetic control brought the pooled estimate to the null. Strong control of clinical indication and social-behavioural factors left residual associations. These results link attenuation most consistently to shared familial liability in the available evidence. Bias-domain triangulation offers a reusable, pre-pooling test of which unresolved bias structure accompanies non-convergent observational estimates. The study was registered on PROSPERO (CRD420261365276).

## Introduction

Causal interpretation is difficult when an exposure and an outcome share several determinants. This problem is acute for autism spectrum disorder (ASD) and attention-deficit/hyperactivity disorder (ADHD), two highly heritable, polygenic neurodevelopmental conditions. Maternal psychiatric liability, socioeconomic context and pregnancy-related clinical indications can influence both medication use and offspring outcomes^1,2^. These factors also shape maternal prescribing and treatment patterns^3^. Meta-analysis may therefore create apparent agreement when studies leave the same pathways uncontrolled. Prenatal paracetamol (acetaminophen) has become a prominent example. A regulatory announcement in September 2025 suggested a possible association with offspring autism and was followed by public concern and litigation^4^. The unresolved question is whether the association reflects a causal effect or differences in control of familial, clinical and behavioural confounding^5^.

Previous syntheses have not resolved this question. An umbrella review rated the available reviews as low or critically low quality and identified overlap and incompletely addressed confounding^6^. Conventional cohorts often report positive associations, whereas sibling comparisons tend to yield estimates closer to the null^7,8^. This contrast has placed sibling designs at the top of an informal evidence hierarchy^9^. Yet sibling comparisons mainly address shared, time-invariant familial factors. They remain susceptible to non-shared indications and social-behavioural factors^10,11^. Study design alone therefore does not show which back-door pathways remain open.

Existing appraisal and synthesis methods offer limited help. ROBINS-E recognizes confounding but relies on qualitative, study-specific judgments that are difficult to scale. Large language models have not reproduced these judgments consistently^12,13^. Standard meta-analysis also rarely treats adjustment structure as a measurable source of heterogeneity^14^. Calls for stronger causal inference in human behaviour have emphasized systematic triangulation across sources of under-control and over-control^15^. Directed acyclic graphs (DAGs) can make these assumptions explicit before pooling^16-18^. We built on the confounder-matrix approach^19^ to quantify control of distinct causal pathways.

We reconstructed the adjustment sets reported over 12 years using the ESC-DAG protocol^20^. The resulting DAG contained 27 displayed nodes and 50 directed edges. We grouped back-door pathways into familial/genetic liability (B1), clinical indication (B2) and social-behavioural context (B3). Each estimate was rated within these domains before meta-analysis. Using this framework (Fig. 1), we then tested two predictions. Estimates should approach the null as control of a responsible bias domain improves, and strong-control estimates should converge if the domains address the same causal association. This design evaluates why the literature remains non-convergent and provides a framework that can be reused in other observational syntheses.

**Figure 1.**
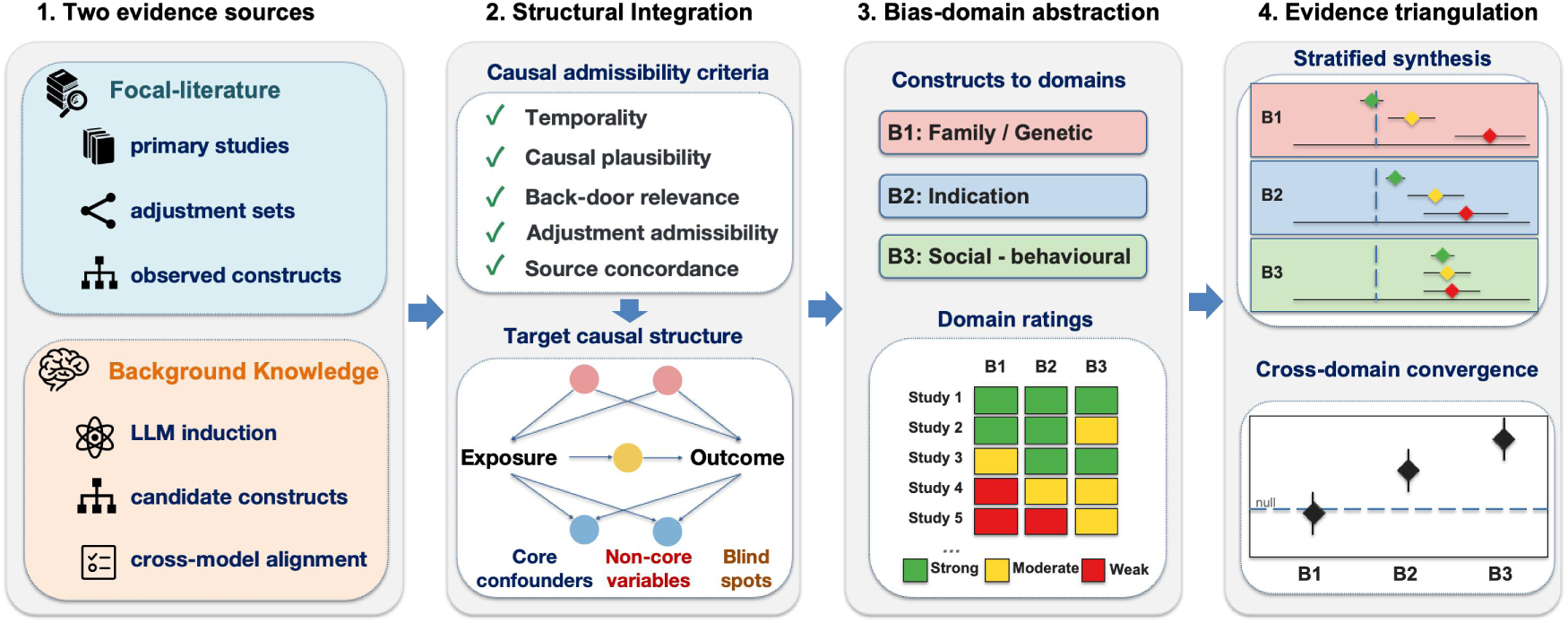
Bias-domain triangulation as a causal-structure-aware evidence synthesis framework. The framework separates two sources of causal information: the focal literature, which records what studies adjusted for, and background knowledge, which generates candidate causal structures that should be considered. These two sources are integrated through causal-role adjudication using pre-specified causality criteria, including temporality, causal plausibility, back-door relevance, adjustment admissibility, and source concordance. The resulting final target DAG is used to define bias domains, exclude non-core variables from confounding-control scores, rate each study within domains, and measure whether existing adjusted estimates converge across bias structures. Convergence across strong-control strata strengthens causal interpretation, whereas non-convergence identifies unresolved bias pathways.

## Results

### Two-source DAG construction

We screened 1,162 unique records identified through MEDLINE and Embase (Supplementary Section 1.2). Of these, 48 articles met the review criteria, and 46 contained enough information for bias-domain classification. A total of 24 articles contributed 39 adjusted ratio-type estimates to the meta-analysis (Supplementary Fig. S1). Collapsing same-design ASD and ADHD estimates within articles yielded 30 article-design combinations^7,21-43^.

We constructed the target DAG before stratified meta-analysis (Fig. 1). Track A recorded the covariates used in each focal study’s main model and harmonised them into constructs. Track B generated candidate determinants, mediators, colliders, selection variables and measurement-related nodes without access to Track A. Across models, Track B produced 1,357 unique candidate variables and three taxonomies of 38, 38 and 27 constructs.

Figure 2 shows the Track B workflow and its validation. Multiple large language models generated candidate variables, which were de-duplicated and grouped into blind construct taxonomies. These taxonomies were then aligned with Track A. Candidate-structure generation was therefore independent of the literature appraisal.

**Figure 2.**
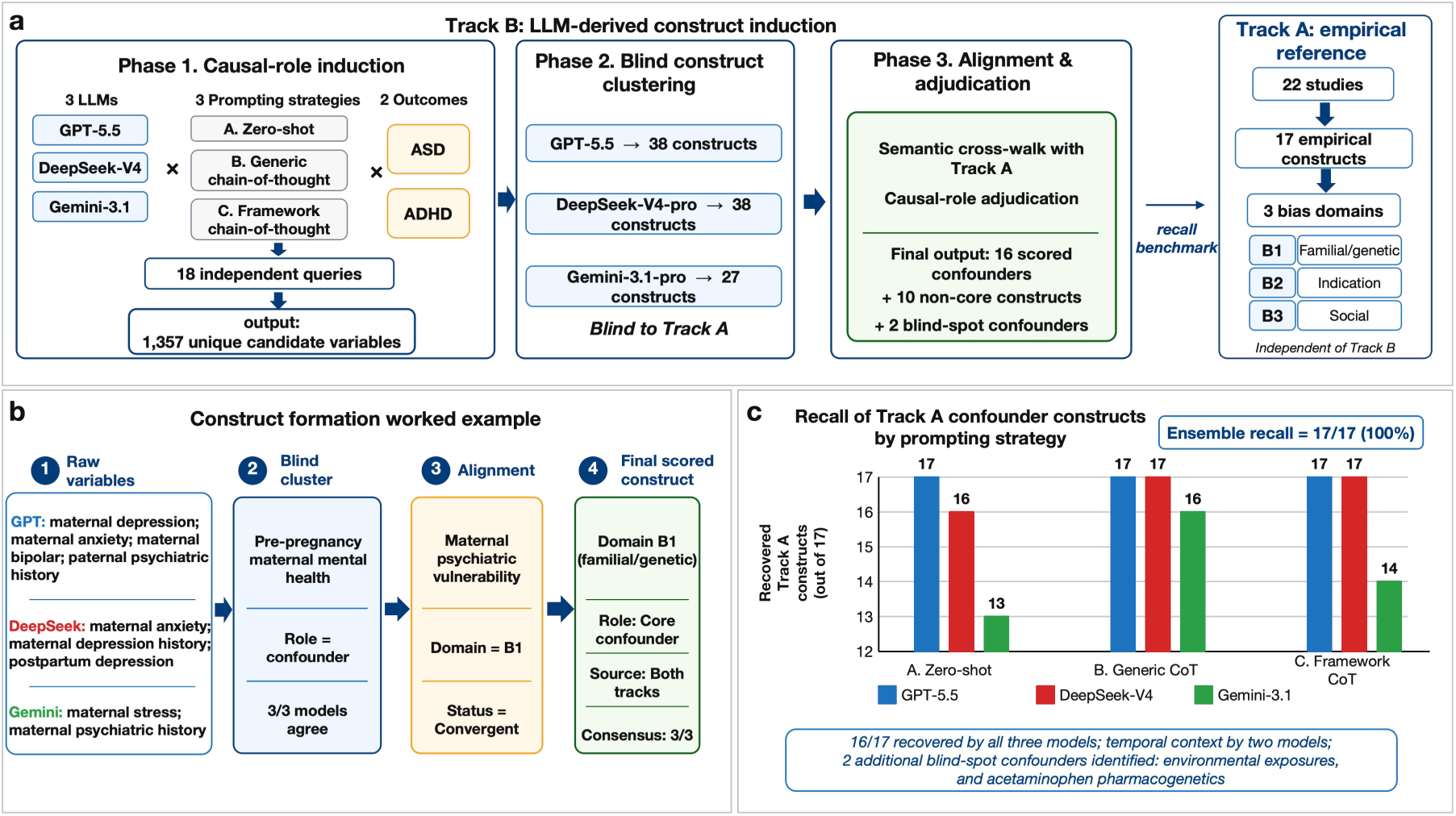
LLM prompting workflow and recall validation for DAG construct generation. a, Track B used three LLMs, three prompting strategies and two outcomes to generate 18 independent queries, yielding 1,357 deduplicated candidate variables. Each model then independently clustered these variables into blind construct taxonomies, which were aligned with the Track A empirical taxonomy derived from 22 focal studies and comprising 17 pre-adjudication Track A constructs. Causal-role adjudication produced 16 scored confounders, 10 non-core constructs and 2 Track-B-only blind-spot confounders. b, Worked example showing how model-generated variables related to maternal psychiatric history were clustered into a Track B construct, aligned with the Track A construct "maternal psychiatric vulnerability", and retained as a core B1 familial/genetic confounder. c, Recall validation showed that the three-model ensemble recovered all 17 pre-adjudication Track A constructs; 16 were recovered by all three models and temporal context by two models. Track B additionally identified blind-spot confounders, including environmental exposures and acetaminophen pharmacogenetics.

The three-model ensemble recovered every Track A construct. Sixteen of 17 constructs were recovered by all three models, and temporal context was recovered by two. Causal-role adjudication reduced the scored set to 16 confounders. Post-exposure pregnancy complications were excluded, whereas pre-exposure obstetric history remained within reproductive history. Track B also identified rarely measured candidate confounders, including environmental or occupational exposures and acetaminophen pharmacogenetics.

The integration used pre-specified criteria for temporality, causal plausibility, back-door relevance, adjustment admissibility and source concordance. The full taxonomy contained 29 conceptual constructs. Sixteen core confounders were scored across B1, B2 and B3. Ten non-core constructs remained visible but were not scored, and three Track B candidate confounders were reported without scoring. Four mechanistic or ascertainment constructs were omitted from, or collapsed within, the displayed graph. The final DAG contained 27 displayed nodes, including exposure and outcome, and 50 directed edges.

Table 1 summarizes the construct set and rating framework. Supplementary Table S1 gives the full mapping and adjudication rationale. Supplementary Table S2 reports recall by model and prompting strategy, and Supplementary Table S3 reports ratings for each estimate.

**Table 1.** Two-source construct set and the bias-domain rating framework.

| Causal category | Constructs included in the final target DAG | Source status | Rating or treatment in analyses |
| --- | --- | --- | --- |
| **B1 familial/genetic liability** | Shared familial/genetic background; maternal neurodevelopmental liability; maternal psychiatric vulnerability | Track A + Track B | Strong = sibling/family fixed-effects; Moderate = $\geq 1$ familial-liability proxy; Weak = none |
| **B2 clinical indication and maternal physical health** | Acute infection/fever; pain/headache/migraine; chronic maternal medical conditions; concomitant medication/treatment context; maternal metabolic/adiposity status | Track A + Track B | Strong = broad B2 coverage including direct indication; Moderate = partial B2 coverage; Weak = none |
| **B3 social-behavioural context** | Socioeconomic position; demographic/regional context; lifestyle/substance use; psychosocial stress; maternal demographic context; reproductive history/parity; family structure; temporal context | Track A + Track B | Strong = broad B3 coverage; Moderate = partial B3 coverage; Weak = none |
| **Non-core post-exposure or selection structures** | Perinatal mediators; current-pregnancy complications; breastfeeding/lactation; early-child infection/microbiome/atopy; early developmental phenotype; mechanistic chain; selection/cohort entry; diagnostic/ascertainment; healthcare-utilization detection; child sex/age at assessment | Track A + Track B or reclassified | Retained for transparency but excluded from confounding-control scores |
| **Reported confounders (not scored)** | Environmental/occupational exposures; acetaminophen pharmacogenetics; medication-seeking propensity | Track B / split | In the DAG; not scored |

### Confounder matrix across studies

Control differed markedly across bias domains (Fig. 3). Only sibling comparisons and family fixed-effects designs achieved strong B1 control. Measured proxies could not fully address latent shared familial and genetic liability. Most article-design combinations achieved moderate or strong control for B2 and B3.

**Figure 3.**
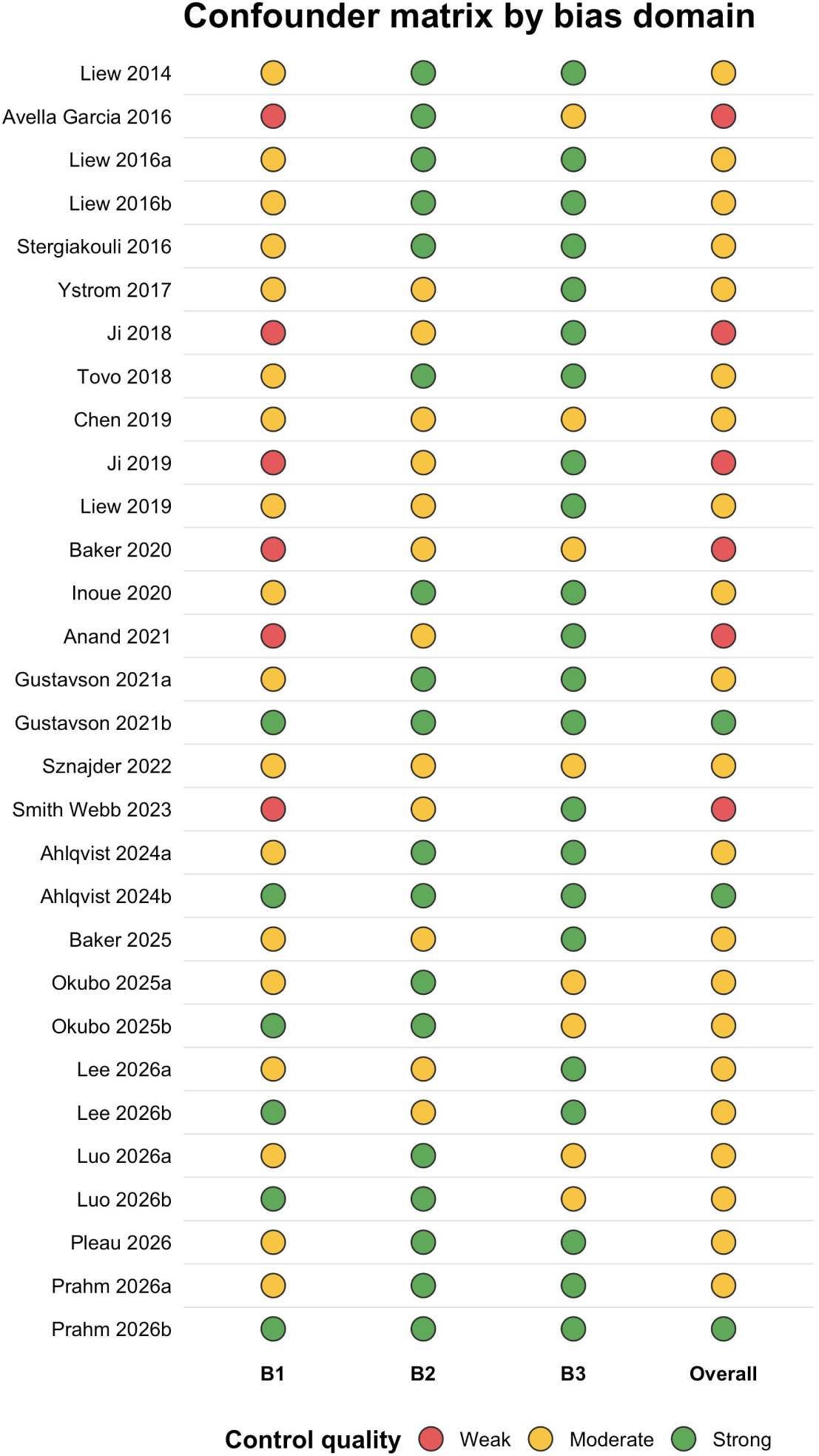
Confounder matrix by bias domain. Confounder matrix for 30 article-design combinations after collapsing same-design ASD and ADHD outcomes within articles. Each circle shows control quality (Strong, Moderate or Weak) for B1 familial/genetic liability, B2 clinical indication and maternal physical health, B3 social-behavioural context and overall control, under the locked 16-construct two-source mapping.

No single study label captured adjustment quality. Studies with strong clinical or social-behavioural control could retain weak or moderate B1 control. Family-based designs addressed B1 but differed in their control of pregnancy-specific indications and social context. Only four estimates achieved strong overall control.

### Stratified meta-analysis by overall confounding control

The pooled association across 39 estimates was 1.15 (95% CI 1.07–1.22). Leave-one-out estimates ranged from 1.13 to 1.15. No individual estimate determined the result. Stratification then tested whether this average concealed systematic differences in confounding control.

Overall control followed a weakest-link rule. Strong ratings required strong control in all three domains, and any weak domain produced a weak overall rating. All other estimates were moderate. Weak-control estimates pooled to 2.08 (95% CI 1.57–2.75; n = 7). Moderate-control estimates pooled to 1.12 (95% CI 1.07–1.17; n = 28). Strong-control estimates pooled to 0.98 (95% CI 0.95–1.02; n = 4). The subgroup-interaction p value was <0.0001 (Supplementary Fig. S5). Greater joint control accompanied attenuation, but the overall score could not identify the responsible domain.

### Domain-specific attenuation of pooled effect estimates

B1 showed a monotonic attenuation gradient. Weak-control estimates pooled to 2.08 (95% CI 1.57–2.75; n = 7). Moderate-control estimates pooled to 1.15 (95% CI 1.10–1.20; n = 22). Strong-control estimates pooled to 0.99 (95% CI 0.97–1.00; n = 10). The subgroup-interaction p value was <0.0001 (Fig. 4). Only sibling comparisons and family fixed-effects designs qualified as strong B1 control.

**Figure 4.**
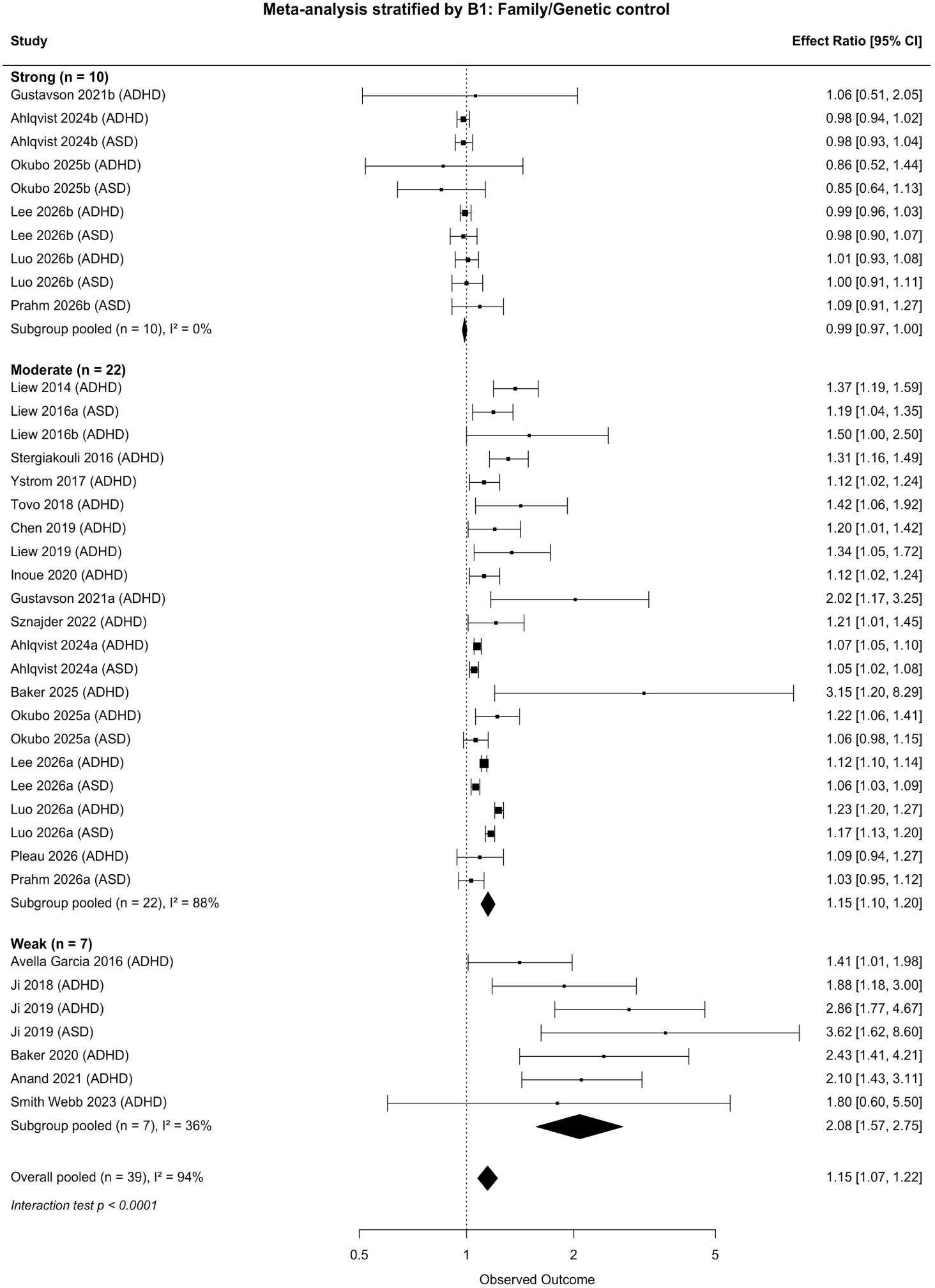
Random-effects meta-analysis of prenatal acetaminophen exposure and offspring ASD/ADHD, stratified by the strength of familial/genetic (B1) confounding control. The pooled association falls monotonically as control improves: 2.08 (1.57–2.75; 7 estimates) under weak control, 1.15 (1.10–1.20; 22) under moderate, and 0.99 (0.97–1.00; 10) under strong control (subgroup-interaction p < 0.0001). Strong B1 control is achieved by sibling-comparison or family-fixed-effects designs. The corresponding forests for B2, B3 and overall control are shown in Supplementary Figures S3 to S5.

B2 showed a smaller, non-significant difference between control levels. Pooled estimates were 1.42 (95% CI 1.14–1.78; n = 15) under moderate control and 1.11 (95% CI 1.05–1.17; n = 24) under strong control. The subgroup-interaction p value was 0.168 (Supplementary Fig. S3). No estimate was rated weak for B2.

B3 showed no attenuation gradient. Pooled estimates were 1.13 (95% CI 1.03–1.24; n = 12) under moderate control and 1.18 (95% CI 1.07–1.30; n = 27) under strong control. The subgroup-interaction p value was 0.752 (Supplementary Fig. S4). No estimate was rated weak for B3.

Thus, attenuation tracked increasing control of B1, but not B2 or B3. Strong-control estimates for B2 and B3 remained above the null.

### Cross-bias-domain convergence

Strong-control estimates did not converge across domains. They were 0.99 for B1, 1.11 for B2 and 1.18 for B3 (Fig. 5d). Only B1 showed attenuation within the domain (Fig. 5a–c). The observed pattern was therefore specific to familial/genetic control in this evidence base.

**Figure 5.**
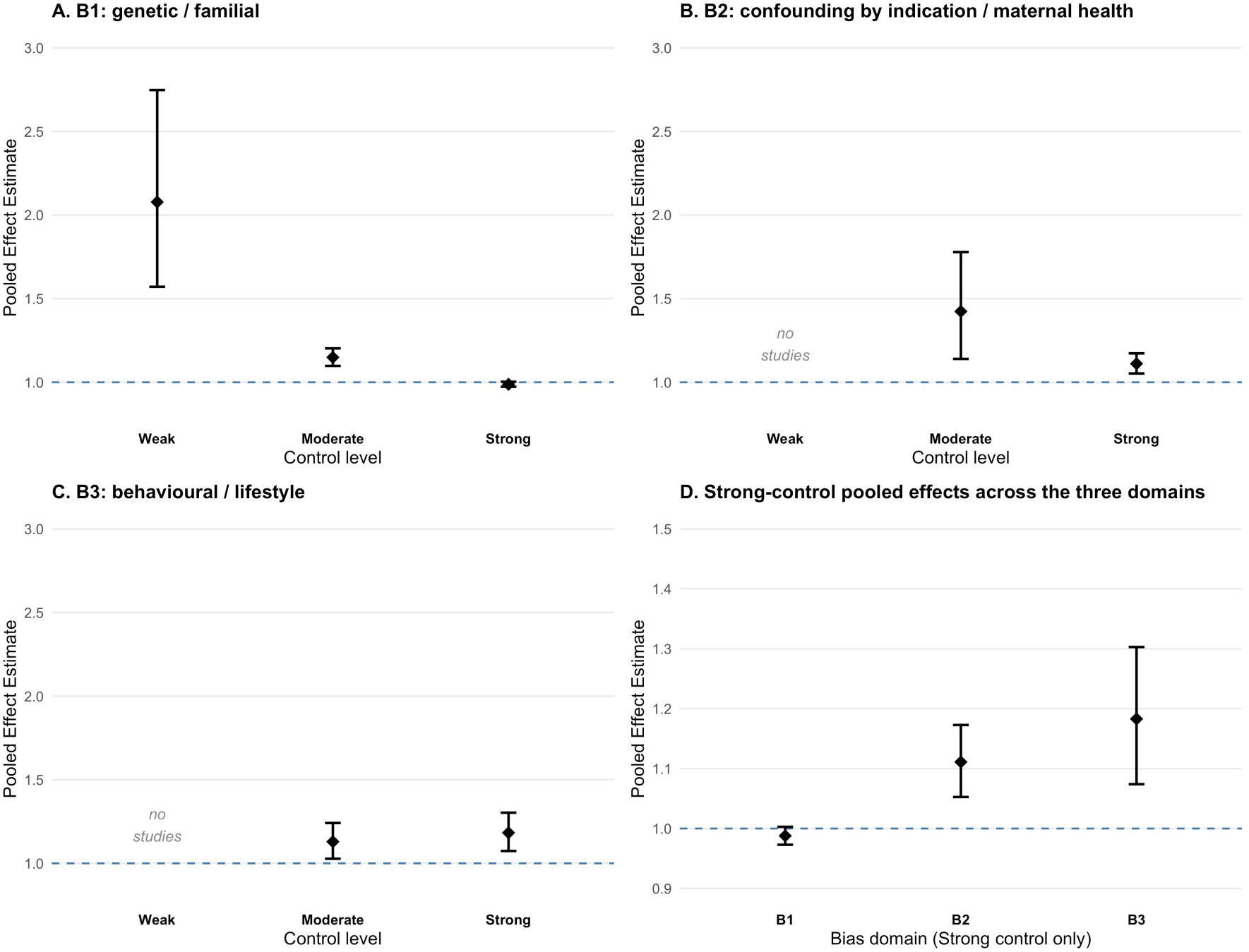
Triangulation dashboard demonstrating within- and cross-domain non-convergence. a–c, Pooled estimate within each bias domain stratified by control strength (Weak / Moderate / Strong): a, B1 genetic/familial (n = 7 / 22 / 10); b, B2 indication/maternal health (n = 0 / 15 / 24); c, B3 sociodemographic/lifestyle (n = 0 / 12 / 27). d, Strong-control pooled estimates across the three domains; only B1 reaches the null.

### Sensitivity analysis

Sensitivity analyses preserved the main pattern. The design-gated B1 gradient was unchanged across construct granularities and rating thresholds. Among 24 HR/aHR estimates, the overall pooled estimate was 1.08 (95% CI 1.03–1.12). B1 estimates pooled to 0.99 (0.97–1.00; n = 10) under strong control and 1.13 (1.07–1.18; n = 14) under moderate control. The subgroup-interaction p value was 0.0004, and no HR/aHR estimate had weak B1 control. Leave-one-out estimates ranged from 1.13 to 1.15. Supplementary Tables S3–S5 and Source Data provide complete results.

## Discussion

We developed bias-domain triangulation to test whether non-convergent estimates track control of specific causal pathways. The framework separates reported adjustment sets from an independent background structure before pooling. In this application, estimates approached the null only under strong familial/genetic control. Strong clinical-indication and social-behavioural control left residual associations. These results connect attenuation most consistently to shared familial liability in the available literature.

This framework extends quantitative evidence triangulation by making adjustment structure the unit of comparison^44,45^. Earlier syntheses reported attenuation in sibling designs^6,8,9^, but the design label does not identify the controlled pathway. Sibling comparisons can address shared familial factors while leaving non-shared indications and social context unresolved. We harmonised hundreds of reported variables into 29 constructs using ESC-DAG^20^. This representation allowed studies to be compared by causal topology rather than design name.

The findings also expose a limitation of ROBINS-E^13^ and ROBINS-I^46^. These tools evaluate confounding within broader risk-of-bias assessments, but the final judgment remains qualitative and study-specific. The matrix instead represents adjustment along several causal dimensions. Family-based designs can control shared, time-invariant causes while retaining non-shared clinical or social confounding. They can also adjust for downstream variables. B2 and B3 did not reproduce the B1 gradient, showing that covariate number alone does not capture adjustment quality.

Explicit causal diagrams were rare in the focal literature, despite widespread covariate adjustment. Without a graph, adjustment may leave back-door paths open, block causal pathways or induce collider bias. This concern accords with critiques of under-control and over-control in mental and behavioural research^15^. Bias-domain mapping assigns each construct a single causal role and prevents cross-domain double counting. Constructing the target DAG before pooling also makes the underlying assumptions available for scrutiny.

The analysis has implications for decisions based on real-world evidence. The September 2025 announcements on prenatal paracetamol show how observational associations can enter policy before major bias structures are resolved. The volume of evidence cannot compensate for shared residual confounding. Structural bias audits could therefore complement conventional evidence grading in pharmacovigilance and guideline development. They can show whether an association persists after control of a specified back-door pathway. Consequences for treatment decisions require direct clinical evaluation and cannot be inferred from this literature-level analysis alone.

Several limitations define the scope of these conclusions. Construct definitions and rating thresholds required expert judgment, although the DAG and criteria were specified explicitly. Familial/genetic liability may remain incompletely controlled outside sibling or family fixed-effects designs. Only four estimates achieved strong overall control, limiting precision. The framework evaluates confounding structure but does not resolve misclassification, selection bias or publication bias. Finally, this single pharmacoepidemiological application cannot establish performance across other exposure-outcome questions.

Bias-domain triangulation converts adjustment structure into a quantitative target for synthesis. In this literature, attenuation followed stronger familial/genetic control but not stronger control in the other domains. The framework identifies which bias pathway accompanies estimate change, while preserving uncertainty about causality and generalisability.

## Online Methods

### Data extraction and verification

Two annotators independently extracted the main-model covariates and pooled estimate from each source full-text PDF. Covariates were mapped blindly to the consolidated constructs before comparison. Exact agreement for effect-size transcription was 78.8% (ICC 0.835, 95% CI 0.690–0.910; Lin’s concordance 0.831). Construct coding yielded Cohen’s κ = 0.847 and 93.4% observed agreement. Discrepancies were resolved by PDF review and senior adjudication. Source Data contain the agreement statistics and record-level decisions.

### DAG construction and bias domain definition

The framework used two sources to define constructs and bias domains. Track A recorded the covariates used by the focal studies. Track B generated an independent background causal structure. This separation prevented the focal literature from defining its own appraisal criteria.

Track A recorded each study’s main adjustment set. Following ESC-DAG^20^, we extracted covariates from the model contributing the meta-analytic estimate. Variables used only in sensitivity, stratified or mediation analyses were excluded. Reported variables were harmonised into constructs using the confounder-matrix approach^19^. Source Data provide the covariate set for each record.

Track B generated candidate background structure without access to Track A. We used OpenAI GPT-5.5, DeepSeek-V4-pro and Google Gemini-3.1-pro-preview. The models came from three independent developers and received neither the Track A taxonomy nor the bias-domain scheme.

Each model evaluated prenatal paracetamol and offspring ASD or ADHD under three prompting strategies. These were zero-shot, generic chain-of-thought and framework chain-of-thought. DeepSeek-V4-pro used temperature 0 for both phases. GPT-5.5 used reasoning_effort=medium without an explicit temperature parameter. Gemini-3.1-pro-preview used temperature 0 for candidate-variable induction and no explicit temperature parameter for blind construct clustering. The models assigned one causal role and a temporal position to every candidate variable.

Variables were pooled across models, prompts and outcomes, then de-duplicated to 1,357 candidates. Each model independently grouped this list while remaining blind to Track A. The resulting taxonomies contained 38, 38 and 27 constructs and separated confounders, mediators and colliders.

The causal-graph terms used in this inspection are defined in Box 1.

#### Box 1. Causal-graph terminology used in this study

We encode the assumed causal structure as a directed acyclic graph (DAG): variables (nodes) joined by arrows (directed edges) with no directed cycle, where an arrow represents a hypothesised direct cause. The exposure node is prenatal acetaminophen use and the outcome node is offspring autism spectrum disorder or attention-deficit/hyperactivity disorder. Because a variable’s position in this graph determines whether adjusting for it removes or introduces bias, we classify each candidate variable by its role:

**Confounder.** A shared cause of the exposure and the outcome (an arrow into each). Adjustment blocks a back-door path and reduces bias; leaving it unadjusted leaves confounding. Confounders are the variables the framework scores.

**Mediator.** A variable lying on a directed path from the exposure to the outcome (an incoming and an outgoing arrow along that path). Adjustment removes part of the effect under study (over-adjustment), so mediators are excluded from scoring.

**Collider.** A shared effect of two or more nodes (two or more incoming arrows). Adjusting for it, or for its descendant, opens a non-causal path and can create bias (collider or M-bias); colliders are excluded.

**Instrument.** A cause of the exposure that affects the outcome only through the exposure. It is not a confounder and is kept out of the adjustment set.

**Latent common cause.** An unmeasured shared cause, for example familial and genetic liability, that cannot be adjusted directly; it is addressed by study design or by an observed proxy.

In terms of node degree, a confounder has outgoing arrows to both the exposure and the outcome, a mediator has an incoming and an outgoing arrow on the exposure-to-outcome path, and a collider has in-degree greater than one. This role taxonomy underlies the variable-role inspection and the bias-domain scoring described below.

The 38-construct taxonomy contained a semantic equivalent of every construct in the other two taxonomies. We therefore used it as the Track B reference. Track B constructs were mapped to Track A as convergent, Track B-only or role-conflicting. Supplementary Table S1 reports the integrated construct set.

Causal roles were adjudicated using pre-specified criteria for temporality, biological plausibility, back-door relevance, adjustment admissibility and source concordance. Pre-exposure common causes were eligible for scoring. Mediators, colliders and precision covariates remained visible but were excluded from the score.

The ensemble recovered all 17 pre-adjudication Track A constructs. One construct, current-pregnancy obstetric complications, was subsequently reclassified as a mediator; the final scored confounder set therefore contained 16 constructs. Sixteen were recovered by all three models, and temporal context by two. Adjudication produced three changes. Pre-exposure obstetric history was retained, whereas current-pregnancy complications were treated as mediators. Environmental or occupational exposures and acetaminophen pharmacogenetics were reported as unscored candidate confounders. Healthcare utilisation was separated into medication-seeking propensity and an ascertainment pathway. Supplementary Table S2 gives recall by model and prompting strategy.

The scoring set contained 16 confounder constructs. B1 included three familial/genetic constructs, B2 included five clinical-indication or maternal-health constructs, and B3 included eight sociodemographic or lifestyle constructs. Non-core constructs remained in the matrix but were not scored. Supplementary Figure S2 shows the consolidated DAG.

### Criteria for confounding control strength

Selection of an adjustment set can follow three established criteria, summarised in Box 2.

#### Box 2. Criteria for selecting the adjustment set

Three broad criteria are used to decide which variables to adjust for.

**Pre-exposure (temporality) criterion.** Adjust for every variable measured before the exposure. It is simple and robust to mis-specification of causal direction, but it may inadvertently include a pre-exposure collider and thereby induce M-bias.

**Common-cause criterion.** Adjust only for variables that are common causes of both the exposure and the outcome. It is theoretically correct and avoids conditioning on colliders, but it requires near-complete causal knowledge and leaves residual confounding when a common cause is unmeasured.

**Modified disjunctive cause criterion.** Adjust for any cause of the exposure, the outcome, or both; exclude instruments (causes of the exposure only); and, where a common cause is unmeasured, include an observed proxy. It is among the most comprehensive practical strategies and mitigates the limitations of the other two, its main difficulty being the identification of instruments.

The present framework applies the common-cause criterion: it scores only pre-exposure common causes of both the exposure and the outcome, and excludes mediators, colliders, instruments, and precision (outcome-only) variables from scoring. It is therefore more restrictive than the disjunctive set. Consistent with the modified disjunctive cause criterion in one respect, an unmeasured common cause (the latent familial and genetic construct, B1) is credited when controlled by study design or an observed proxy rather than left unaddressed.

Ratings represented the number of back-door pathways addressed, not the raw number of covariates. Adjustment for an adequate proxy counted as control of the corresponding construct. Strong B2 control required at least three of five constructs, and moderate control required one or two. Strong B3 control required at least four of eight constructs, and moderate control required one to three. No adjusted construct produced a weak rating in either domain.

B1 used a design-gated rule because shared familial/genetic background is latent. Strong control required a sibling comparison or family fixed-effects design. Population analyses were moderate if they adjusted for maternal neurodevelopmental traits or psychiatric vulnerability. Analyses without either proxy were weak.

Domain ratings were strong, moderate or weak. Overall control followed a weakest-link rule. Strong required all three domains to be strong, and weak required at least one weak domain. All other combinations were moderate.

### Statistical analysis

Hazard, risk and odds ratios were pooled on the log scale using random-effects models. We used REML estimation with Knapp–Hartung inference and a categorical moderator Q-test. Sensitivity analyses varied the rating thresholds and used taxonomies with 11, 16 or 19 confounder constructs. We also restricted the data to HR/aHR estimates and performed leave-one-out analyses.

## Data & Code availability

All data supporting the findings of this study are available in the article, its Supplementary Information and Source Data files. No individual-level participant data were used; all extractions are from published study reports. All analysis code, including R scripts for the stratified meta-analysis, Python scripts for bias-domain rating and LLM-based DAG induction, and the exact prompts sent to the three large language models, are available at https://github.com/xuanyshi/bias-domain-triangulation. A step-by-step protocol for applying the framework to other exposure– outcome questions is provided in the Supplementary Information and as an interactive web tutorial at https://xuanyshi.github.io/bias-domain-triangulation/.

## Data Availability

No individual-level participant data were used; all extractions are from published study reports. All analysis code, including R scripts for the stratified meta-analysis, Python scripts for bias-domain rating and LLM-based DAG induction, and the exact prompts sent to the three large language models, are available at https://github.com/xuanyshi/bias-domain-triangulation.

## Acknowledgements

This work is supported by Michigan Medicine-PKUHSC Joint Institute (JI) Award, (BMU2026JIUM001) and the National Natural Science Foundation of China (82330107).

## Author contributions

Conceptualization: J.D., X.S. Methodology: J.D., X.S. Formal analysis: X.S. Data Curation: X.S., G.D. Writing, original draft: X.S., J.D. Writing, review and editing: X.S., J.D. Funding acquisition: J.D.

## References

1. Tick, B., Bolton, P., Happé, F., Rutter, M. & Rijsdijk, F. Heritability of autism spectrum disorders: a meta-analysis of twin studies. Journal of Child Psychology and Psychiatry 57, 585–595 (2016).

2. Faraone, S.V. & Larsson, H. Genetics of attention deficit hyperactivity disorder. Molecular psychiatry 24, 562–575 (2019).

3. Mansour, O., et al. Prescription medication use during pregnancy in the United States from 2011 to 2020: trends and safety evidence. American journal of obstetrics and gynecology 231, 250.e251–250.e216 (2024).

4. Lang, K. Trump’s claims on Tylenol (paracetamol), vaccines, and autism—what’s the truth? BMJ (Clinical research ed.) 390, r2025 (2025).

5. Prada, D., Ritz, B., Bauer, A.Z. & Baccarelli, A.A. Evaluation of the evidence on acetaminophen use and neurodevelopmental disorders using the Navigation Guide methodology. Environmental health : a global access science source 24, 56 (2025).

6. Sheikh, J., et al. Maternal paracetamol (acetaminophen) use during pregnancy and risk of autism spectrum disorder and attention deficit/hyperactivity disorder in offspring: umbrella review of systematic reviews. BMJ (Clinical research ed.) 391, e088141 (2025).

7. Ahlqvist, V.H., et al. Acetaminophen Use During Pregnancy and Children’s Risk of Autism, ADHD, and Intellectual Disability. Jama 331, 1205–1214 (2024).

8. D’Antonio, F., et al. Prenatal paracetamol exposure and child neurodevelopment: a systematic review and meta-analysis. *The Lancet Obstetrics, Gynaecology*, & Women’s Health 2, e190–e198 (2026).

9. Masarwa, R., et al. Prenatal Exposure to Acetaminophen and Risk for Attention Deficit Hyperactivity Disorder and Autistic Spectrum Disorder: A Systematic Review, Meta-Analysis, and Meta-Regression Analysis of Cohort Studies. American journal of epidemiology 187, 1817–1827 (2018).

10. Masarwa, R., Platt, R.W. & Filion, K.B. Acetaminophen use during pregnancy and the risk of attention deficit hyperactivity disorder: A causal association or bias? Paediatric and perinatal epidemiology 34, 309–317 (2020).

11. Ricci, C., et al. In utero acetaminophen exposure and child neurodevelopmental outcomes: Systematic review and meta-analysis. Paediatric and perinatal epidemiology 37, 473–484 (2023).

12. Hasan, B., et al. Integrating large language models in systematic reviews: a framework and case study using ROBINS-I for risk of bias assessment. BMJ evidence-based medicine 29, 394–398 (2024).

13. Higgins, J.P.T., et al. A tool to assess risk of bias in non-randomized follow-up studies of exposure effects (ROBINS-E). Environment international 186, 108602 (2024).

14. Rijnhart, J.J.M., Rabbers, A., Rizzuto, S., Howard, A.M. & Valente, M.J. An umbrella review reveals that control variables are rarely considered as a source of heterogeneity in systematic reviews of observational studies. Journal of clinical epidemiology 184, 111826 (2025).

15. Bailey, D.H., et al. Causal inference on human behaviour. Nature human behaviour 8, 1448–1459 (2024).

16. Dijk, S.W., Caulley, L.M., Hunink, M. & Labrecque, J. From complexity to clarity: how directed acyclic graphs enhance the study design of systematic reviews and meta-analyses. European journal of epidemiology 39, 27–33 (2024).

17. McIntyre, K.J., Tassiopoulos, K.N., Jeffrey, C., Stranges, S. & Martin, J. Using causal diagrams within the Grading of Recommendations, Assessment, Development and Evaluation framework to evaluate confounding adjustment in observational studies. Journal of clinical epidemiology 175, 111532 (2024).

18. Deng, G. & Du, J. Construction and Application of Directed Acyclic Graphs in Leading Medical Journals. JAMA network open 9, e2553803 (2026).

19. Petersen, J.M., et al. The confounder matrix: A tool to assess confounding bias in systematic reviews of observational studies of etiology. Research synthesis methods 13, 242–254 (2022).

20. Ferguson, K.D., et al. Evidence synthesis for constructing directed acyclic graphs (ESC-DAGs): a novel and systematic method for building directed acyclic graphs. International journal of epidemiology 49, 322–329 (2020).

21. Liew, Z., Ritz, B., Rebordosa, C., Lee, P.-C. & Olsen, J. Acetaminophen use during pregnancy, behavioral problems, and hyperkinetic disorders. JAMA pediatrics 168, 313–320 (2014).

22. Avella-Garcia, C.B., et al. Acetaminophen use in pregnancy and neurodevelopment: attention function and autism spectrum symptoms. International journal of epidemiology 45, 1987–1996 (2016).

23. Liew, Z., Ritz, B., Virk, J. & Olsen, J. Maternal use of acetaminophen during pregnancy and risk of autism spectrum disorders in childhood: AD anish national birth cohort study. Autism Research 9, 951–958 (2016).

24. Liew, Z., Bach, C.C., Asarnow, R.F., Ritz, B. & Olsen, J. Paracetamol use during pregnancy and attention and executive function in offspring at age 5 years. International journal of epidemiology 45, 2009–2017 (2016).

25. Stergiakouli, E., Thapar, A. & Davey Smith, G. Association of acetaminophen use during pregnancy with behavioral problems in childhood: evidence against confounding. JAMA pediatrics 170, 964–970 (2016).

26. Ystrom, E., et al. Prenatal exposure to acetaminophen and risk of ADHD. Pediatrics 140, e20163840 (2017).

27. Ji, Y., et al. Maternal biomarkers of acetaminophen use and offspring attention deficit hyperactivity disorder. Brain sciences 8, 127 (2018).

28. Tovo-Rodrigues, L., et al. Is intrauterine exposure to acetaminophen associated with emotional and hyperactivity problems during childhood? Findings from the 2004 Pelotas birth cohort. BMC psychiatry 18, 368 (2018).

29. Liew, Z., et al. Use of negative control exposure analysis to evaluate confounding: an example of acetaminophen exposure and attention-deficit/hyperactivity disorder in Nurses’ Health Study II. American journal of epidemiology 188, 768–775 (2019).

30. Chen, M.-H., et al. Prenatal Exposure to Acetaminophen and the Risk of Attention-Deficit/Hyperactivity Disorder: A Nationwide Study in Taiwan. The Journal of clinical psychiatry 80, 18m12612–12618m12612 (2019).

31. Ji, Y., et al. Association of cord plasma biomarkers of in utero acetaminophen exposure with risk of attention-deficit/hyperactivity disorder and autism spectrum disorder in childhood. JAMA psychiatry 77, 180–189 (2020).

32. Inoue, K., et al. Behavioral problems at age 11 years after prenatal and postnatal exposure to acetaminophen: parent-reported and self-reported outcomes. American journal of epidemiology 190, 1009–1020 (2021).

33. Baker, B.H., et al. Association of prenatal acetaminophen exposure measured in meconium with risk of attention-deficit/hyperactivity disorder mediated by frontoparietal network brain connectivity. JAMA pediatrics 174, 1073–1081 (2020).

34. Anand, N.S., et al. Maternal and cord plasma branched-chain amino acids and child risk of attention - deficit hyperactivity disorder: a prospective birth cohort study. Journal of Child Psychology and Psychiatry 62, 868–875 (2021).

35. Gustavson, K., et al. Acetaminophen use during pregnancy and offspring attention deficit hyperactivity disorder–a longitudinal sibling control study. JCPP advances 1, e12020 (2021).

36. Sznajder, K.K., Teti, D.M. & Kjerulff, K.H. Maternal use of acetaminophen during pregnancy and neurobehavioral problems in offspring at 3 years: A prospective cohort study. PLoS One 17, e0272593 (2022).

37. Smith-Webb, R.S., Barnard-Mayers, R., Werler, M.M. & Parker, S.E. Prenatal exposure to acetaminophen and adolescent assessment of behavior: discrepancies by age and reporter. Frontiers in pharmacology 14, 1084781 (2023).

38. Okubo, Y., Hayakawa, I., Sugitate, R. & Nariai, H. Maternal acetaminophen use and offspring’s neurodevelopmental outcome: a nationwide birth cohort study. Paediatric and perinatal epidemiology 40, 70–79 (2026).

39. Baker, B.H., et al. Associations of maternal blood biomarkers of prenatal APAP exposure with placental gene expression and child attention deficit hyperactivity disorder. Nature Mental Health 3, 318–331 (2025).

40. Pleau, J., Leal, L.F., Sheehy, O. & Bérard, A. The debate on acetaminophen use in pregnancy and neurodevelopmental disorders: Facts or fiction? Journal of Obstetrics and Gynaecology Canada, 103176 (2025).

41. Lee, P.-C., et al. Maternal Acetaminophen Use and Child Neurodevelopment. JAMA pediatrics (2026).

42. Prahm, K.P., et al. Acetaminophen Exposure During Pregnancy and the Risk of Autism in Offspring. JAMA Pediatrics 180, 687–690 (2026).

43. Luo, S., et al. Prenatal acetaminophen (paracetamol) use and the risk of autism and/or attention-deficit/hyperactivity disorder among sibling-matched cohorts. JAMA Internal Medicine (2026).

44. Shi, X., Zhao, W., Chen, T., Yang, C. & Du, J. Evidence triangulator: using large language models to extract and synthesize causal evidence across study designs. Nature communications 16, 7355 (2025).

45. Gutierrez, S., Glymour, M.M. & Smith, G.D. Evidence triangulation in health research. European journal of epidemiology 40, 743–757 (2025).

46. Sterne, J.A., et al. ROBINS-I: a tool for assessing risk of bias in non-randomised studies of interventions. BMJ (Clinical research ed.) 355, i4919 (2016).

