## Supplementary information for "Bias-domain triangulation of non-convergent observational evidence in mental health research"

This Supplement covers study selection, the two-source construction of the consolidated directed acyclic graph (DAG) and bias domains, the construct set, the per-estimate ratings and sensitivity analyses, and the Track B large-language-model prompts. Supplementary Tables are numbered S1 to S7 and Supplementary Figures S1 to S5.

#### 1. Study selection

##### 1.1 Eligibility criteria

Inclusion criteria:

- Design: observational studies (prospective or retrospective cohort, sibling-comparison or other family-based designs).
- Population: pregnant women and their liveborn offspring.
- Exposure: maternal acetaminophen (paracetamol) use during pregnancy.
- Comparator: offspring not exposed, or exposed at a lower level.
- Outcome: offspring autism spectrum disorder and/or attention-deficit/hyperactivity disorder, ascertained by clinical diagnosis, registry linkage or validated instruments.
- Estimates: reported an adjusted ratio-type effect estimate (hazard, odds or rate ratio) with a measure of uncertainty, or provided data from which one could be derived (applied when identifying the studies contributing to the quantitative synthesis).
- Human studies, with no language or publication-date restriction.

Exclusion criteria:

- Reviews, meta-analyses, editorials, commentaries and conference abstracts without primary data.
- Studies without a relevant neurodevelopmental outcome (autism spectrum disorder or attention-deficit/hyperactivity disorder).
- Case-control designs.
- Studies reporting no adjusted effect estimate.
- Duplicate or companion reports of the same analysis; the most complete report was retained.
- Reports with insufficient data for extraction.
- Studies restricted to non-human animals.

##### 1.2 Information sources and search strategy

We searched MEDLINE (via PubMed) and Embase (via embase.com) from inception to 1 July 2026, with no language restriction. The search combined three concept blocks (acetaminophen/paracetamol exposure; the pregnancy, prenatal or perinatal period; and neurodevelopmental outcomes) with the Boolean operator AND, each expanded with controlled vocabulary and free-text title and abstract terms. Animal-only records were excluded where the database interface permitted. Records were combined and de-duplicated by PubMed identifier, DOI and normalised title. The full search strings and the number of records returned at each step are given below.

***MEDLINE (via PubMed)***

| Step | Search terms | Results |
|---|---|---|
| #1 | ("acetaminophen"[MeSH Terms]) OR ("acetaminophen"[tiab]) OR ("paracetamol"[tiab]) OR ("N-acetyl-p-aminophenol"[tiab]) OR ("APAP"[tiab]) OR ("Tylenol"[tiab]) | 38,819 |
| #2 | ("pregnancy"[MeSH Terms]) OR ("prenatal exposure delayed effects"[MeSH Terms]) OR ("maternal exposure"[MeSH Terms]) OR ("pregnan*"[tiab]) OR ("prenatal"[tiab]) OR ("antenatal"[tiab]) OR ("perinatal"[tiab]) OR ("in utero"[tiab]) OR ("maternal"[tiab]) OR ("gestation*"[tiab]) | 1,499,094 |
| #3 | ("attention deficit disorder with hyperactivity"[MeSH Terms]) OR ("autism spectrum disorder"[MeSH Terms]) OR ("neurodevelopmental disorders"[MeSH Terms]) OR ("ADHD"[tiab]) OR ("attention deficit"[tiab]) OR ("hyperactiv*"[tiab]) OR ("autism"[tiab]) OR ("autistic"[tiab]) OR ("ASD"[tiab]) OR ("neurodevelop*"[tiab]) | 374,351 |
| #4 | #1 AND #2 AND #3 | 229 |
| #5 | #4 NOT ("animals"[MeSH Terms] NOT "humans"[MeSH Terms]) | 213 |

***Embase (via embase.com)***

| Step | Search terms | Results |
|---|---|---|
| #1 | 'paracetamol'/exp OR 'acetaminophen':ti,ab OR 'paracetamol':ti,ab OR 'n-acetyl-p-aminophenol':ti,ab OR 'apap':ti,ab OR 'tylenol':ti,ab | 144,268 |
| #2 | 'pregnancy'/exp OR 'prenatal exposure'/exp OR 'maternal exposure'/exp OR pregnan*:ti,ab OR prenatal:ti,ab OR antenatal:ti,ab OR perinatal:ti,ab OR 'in utero':ti,ab OR maternal:ti,ab OR gestation*:ti,ab | 1,769,316 |
| #3 | 'attention deficit disorder'/exp OR 'autism'/exp OR 'neurodevelopmental disorder'/exp OR adhd:ti,ab OR 'attention deficit':ti,ab OR hyperactiv*:ti,ab OR autism:ti,ab OR autistic:ti,ab OR asd:ti,ab OR neurodevelop*:ti,ab | 3,597,263 |
| #4 | #1 AND #2 AND #3 | 1,165 |
| #5 | #4 NOT ([animals]/lim NOT [humans]/lim) | 1,142 |

*MeSH, Medical Subject Headings; tiab/ti,ab, title and abstract; exp, explode. Both databases were searched from inception through 1 July 2026 with no language restriction. Combining PubMed (n = 213) and Embase (n = 1,142) yielded 1,355 records; after removal of 193 duplicates, 1,162 unique records were screened. The systematic review included 48 articles, 46 with sufficient information for bias-domain classification, and 24 articles contributing 39 estimates to the quantitative synthesis.*

Figure S1 shows the PRISMA flow for study identification, screening, eligibility and inclusion.

**Figure S1 | PRISMA flow diagram for study identification, screening, and inclusion.**

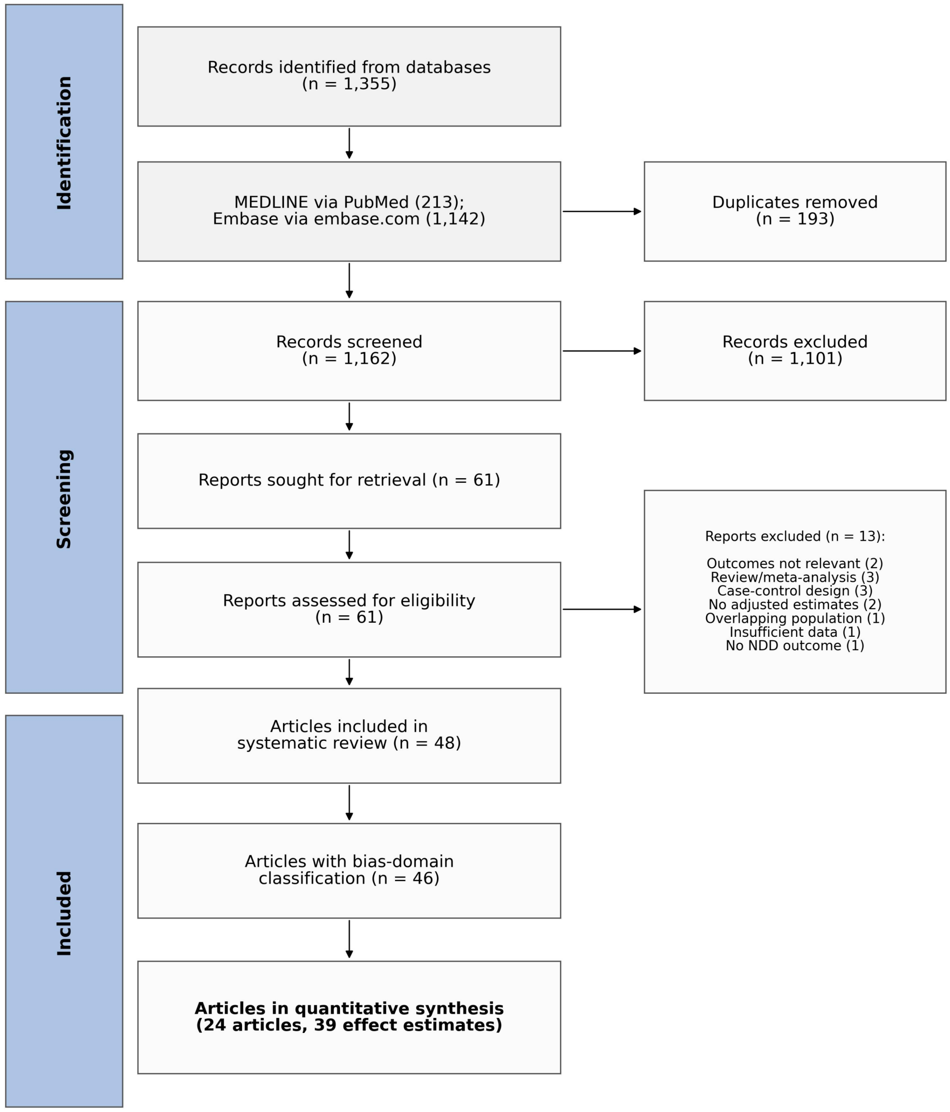

#### 2. Two-source construction of the consolidated DAG

The consolidated DAG and bias domains were built from two independent sources: an empirical audit of study adjustment sets (Track A) and an independent large-language-model induction of the background causal structure (Track B; OpenAI GPT-5.5, DeepSeek-V4-pro, Google Gemini-3.1-pro-preview, three prompting strategies each, blind to Track A). DeepSeek-V4-pro used temperature 0 for both phases. GPT-5.5 used reasoning_effort=medium without an explicit temperature parameter. Gemini-3.1-pro-preview used temperature 0 for candidate-variable induction and no explicit temperature parameter for blind construct clustering. The full procedure is given in the Online Methods. Track B pooled to 1,357 unique candidate variables and three independent taxonomies (38/38/27 constructs); the 38-construct spine was verified to contain a semantic equivalent of every construct in the other two.

**Figure S2 | Consolidated directed acyclic graph (DAG) for prenatal acetaminophen and offspring ASD/ADHD, integrating the Track A empirical audit and the Track B induction (node labels as in Table S1). Confounders (top) comprise the 16 scored constructs across the three bias domains (B1 familial/genetic, B2 indication/maternal health, B3 sociodemographic/lifestyle) together with three Track-B-identified reported confounders drawn as latent (environmental/occupational exposures, acetaminophen pharmacogenetics and medication-seeking propensity). Mediators lie on the exposure-to-outcome path (middle) and colliders are common effects of the exposure and the outcome (bottom); these, with the precision variables, are displayed but excluded from confounding-control scoring.**

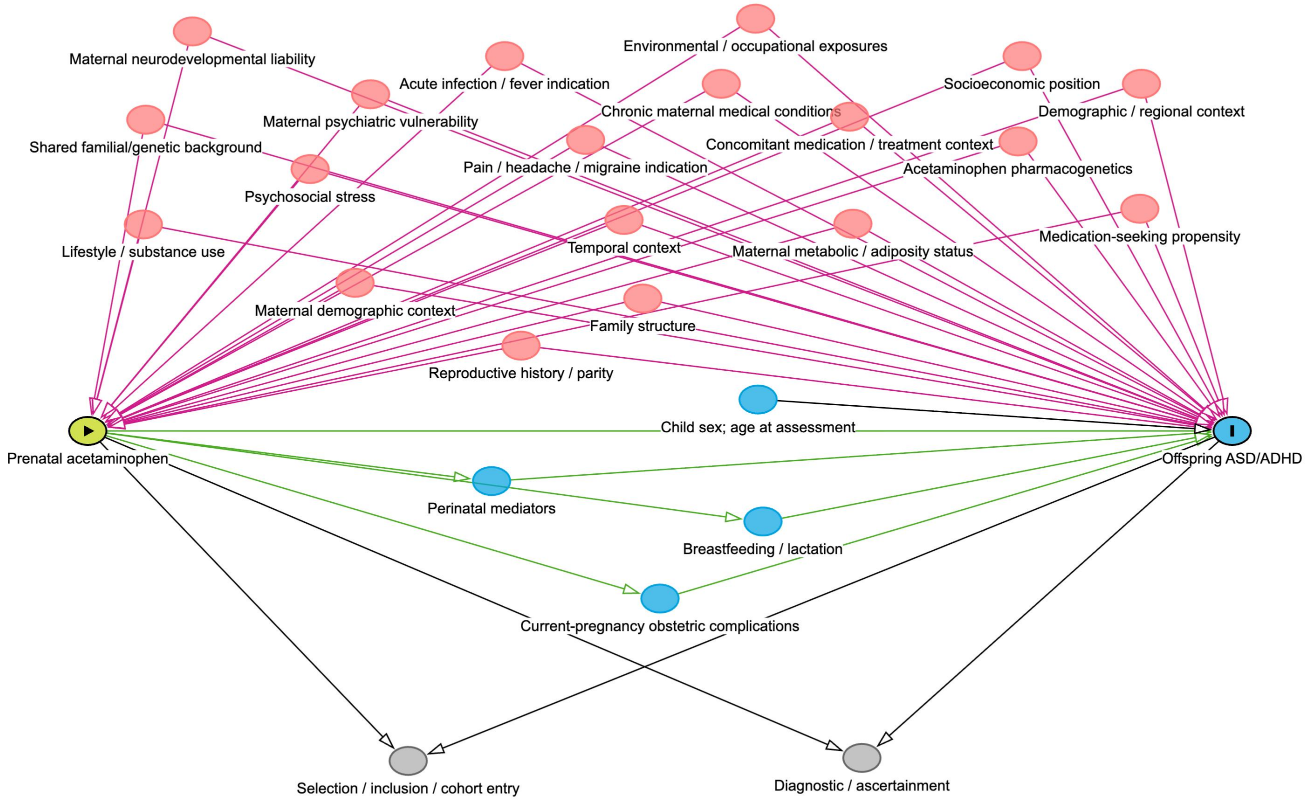

#### 3. Two-source construct set

Table S1 gives the final two-source construct set: each construct's bias domain, adjudicated causal role, source/status, the three-model consensus, whether it is scored, and the over-adjustment concern. Of the 16 scored confounders, 15 were recovered by all three models and one (temporal context) by two. Among the two de novo Track-B-only blind-spot confounders, environmental/occupational exposures were recovered by all three models and acetaminophen pharmacogenetics by two at variable level; medication-seeking propensity was recovered by all three models after the healthcare-utilisation construct was split.

**Table S1 | Final two-source construct set, causal-role adjudication, and bias-domain mapping.**

| Construct | Domain | Causal role | Source / status | 3-model consensus | Over-adj. | Rationale |
|---|---|---|---|---|---|---|
| ① Core confounder constructs, scored (n = 16) | | | | | | |
| Shared familial/genetic background | B1 | Confounder | Both (convergent) | (3/3) | Low | Latent shared-familial back-door; design-gated |
| Maternal neurodevelopmental liability | B1 | Confounder | Both (convergent) | (3/3) | Low | B1 proxy (maternal ADHD/ASD traits) |
| Maternal psychiatric vulnerability | B1 | Confounder | Both (convergent) | (3/3) | Low | B1 proxy (depression/anxiety; shared heritable variance) |
| Acute infection / fever indication | B2 | Confounder | Both (convergent) | (3/3) | Low | Indication common cause |
| Pain / headache / migraine indication | B2 | Confounder | Both (convergent) | (3/3) | Low | Indication common cause |
| Chronic maternal medical conditions | B2 | Confounder | Both (convergent) | (3/3) | Low | Pre-existing maternal disease |
| Concomitant medication / treatment context | B2 | Confounder | Both (convergent) | (3/3) | Low | Co-medication / treatment context |
| Maternal metabolic / adiposity status | B2 | Confounder | Both (convergent) | (3/3) | Low | BMI / diabetes / metabolic |
| Socioeconomic position | B3 | Confounder | Both (convergent) | (3/3) | Low | SES / education / income |
| Demographic / regional context | B3 | Confounder | Both (convergent) | (3/3) | Low | Race / ethnicity / region |
| Lifestyle / substance use | B3 | Confounder | Both (convergent) | (3/3) | Low | Smoking / alcohol / substance |
| Psychosocial stress | B3 | Confounder | Both (convergent) | (3/3) | Low | Prenatal stress / adversity |
| Maternal demographic context | B3 | Confounder | Both (convergent) | (3/3) | Low | Maternal / paternal age |
| Reproductive history / parity | B3 | Confounder | Both (convergent) | (3/3) | Low | Parity; absorbs pre-exposure obstetric history |
| Family structure | B3 | Confounder | Both (convergent) | (3/3) | Low | Marital / partnership context |
| Temporal context | B3 | Confounder | Both (convergent) | (2/3) | Low | Birth year / calendar period |
| ② Non-core constructs, retained in the matrix but excluded from scoring (n = 10) | | | | | | |
| Perinatal mediators (GA, preterm, birth weight, delivery mode) | non-core | Mediator | Both (convergent) | (3/3) | High | Blocks APAP→outcome path |
| Current-pregnancy obstetric complications (preeclampsia/GDM/haemorrhage) | non-core | Mediator | Reclassified | (3/3) | High | Arises in/after exposure window; moved from B2 |
| Breastfeeding / lactation | non-core | Mediator | Both (convergent) | (3/3) | High | Postnatal; blocks part of path |
| Early-child infection / microbiome / atopy | non-core | Mediator | Reclassified | (3/3) | High | Post-birth; relabelled from child characteristic |
| Early developmental / regulatory phenotype | non-core | Mediator | Reclassified | (3/3) | High | Early form of the outcome; never adjust |
| Mechanistic chain (oxidative stress, | non-core | Mediator | Track B | (3/3) | High | Background biological |

| | | | | | | |
|---|---|---|---|---|---|---|
| inflammation, hormonal, epigenetic, placental, fetal brain) | | | (articulated) | | | pathway; post-exposure |
| Selection / inclusion / cohort entry | non-core | Collider | Both (convergent) | (3/3) | High | Conditioning induces collider/M-bias |
| Diagnostic / ascertainment | non-core | Collider | Both (convergent) | (3/3) | High | Descendant of outcome; detection |
| Healthcare utilization → detection | non-core | Collider | Reclassified | (3/3) | High | Ascertainment half of healthcare construct |
| Child sex; age at assessment | non-core | Precision | Both (convergent) | n/a | Low | Precision / measurement-timing, not a confounder |
| ③ Track B-identified confounders, reported but not scored (n = 3) | | | | | | |
| Environmental / occupational exposures (air pollution, pesticides, lead) | Blind-spot | Confounder | Track B (3/3 labs) | (3/3) | n/a | Systematically unadjusted; reported, not scored |
| Acetaminophen pharmacogenetics (GSTT1/GSTM1, CYP2E1) | Blind-spot | Confounder | Track B (2/3 models) | (2/3) | n/a | Only purely-new confounder; unmeasured; reported |
| Medication-seeking propensity | Blind-spot | Confounder | Reclassified | (3/3) | n/a | Confounder half; overlaps B2/B3; unmeasured; reported |

Table S2 reports the variable-level recall underlying this correspondence: for each pre-adjudication Track A construct, the variables that each model elicited under each prompting strategy (A, zero-shot; B, generic chain-of-thought; C, framework chain-of-thought), pooled across outcomes. All 17 pre-adjudication Track A constructs were recovered by the three-model ensemble; 16 were recovered by all three models and one (temporal context) by two. One construct, current-pregnancy obstetric complications, was subsequently reclassified as a mediator; the final scored confounder set therefore contained 16 constructs. Of the two Track B-only blind-spot confounders, environmental/occupational exposures were recovered by all three models and acetaminophen pharmacogenetics by two (gemini surfaced it only at the clustering stage).

**Supplementary Table S2 | Per-model, per-prompting strategy variable-level recall of the Track A confounder constructs (provided as a separate spreadsheet, Supplementary_Table_S2_LLM_DAG_recall.xlsx).**

### 4. Bias-domain ratings and sensitivity analyses

**Table S3 | Per-estimate bias-domain control ratings under the locked mapping.**

| Article/design | Outcome | Effect [95% CI] | B1 | B2 | B3 | Overall |
|---|---|---|---|---|---|---|
| Ahlqvist 2024a | ADHD | 1.07 [1.05–1.10] | Moderate | Strong | Strong | Moderate |
| Ahlqvist 2024a | ASD | 1.05 [1.02–1.08] | Moderate | Strong | Strong | Moderate |
| Ahlqvist 2024b | ADHD | 0.98 [0.94–1.02] | Strong | Strong | Strong | Strong |
| Ahlqvist 2024b | ASD | 0.98 [0.93–1.04] | Strong | Strong | Strong | Strong |
| Anand 2021 | ADHD | 2.10 [1.43–3.11] | Weak | Moderate | Strong | Weak |
| Avella Garcia 2016 | ADHD | 1.41 [1.01–1.98] | Weak | Strong | Moderate | Weak |
| Baker 2020 | ADHD | 2.43 [1.41–4.21] | Weak | Moderate | Moderate | Weak |
| Baker 2025 | ADHD | 3.15 [1.20–8.29] | Moderate | Moderate | Strong | Moderate |
| Chen 2019 | ADHD | 1.20 [1.01–1.42] | Moderate | Moderate | Moderate | Moderate |
| Gustavson 2021a | ADHD | 2.02 [1.17–3.25] | Moderate | Strong | Strong | Moderate |
| Gustavson 2021b | ADHD | 1.06 [0.51–2.05] | Strong | Strong | Strong | Strong |
| Inoue 2020 | ADHD | 1.12 [1.02–1.24] | Moderate | Strong | Strong | Moderate |

| Article/design | Outcome | Effect [95% CI] | B1 | B2 | B3 | Overall |
|---|---|---|---|---|---|---|
| Ji 2018 | ADHD | 1.88 [1.18–3.00] | Weak | Moderate | Strong | Weak |
| Ji 2019 | ADHD | 2.86 [1.77–4.67] | Weak | Moderate | Strong | Weak |
| Ji 2019 | ASD | 3.62 [1.62–8.60] | Weak | Moderate | Strong | Weak |
| Lee 2026a | ADHD | 1.12 [1.10–1.14] | Moderate | Moderate | Strong | Moderate |
| Lee 2026a | ASD | 1.06 [1.03–1.09] | Moderate | Moderate | Strong | Moderate |
| Lee 2026b | ADHD | 0.99 [0.96–1.03] | Strong | Moderate | Strong | Moderate |
| Lee 2026b | ASD | 0.98 [0.90–1.07] | Strong | Moderate | Strong | Moderate |
| Liew 2014 | ADHD | 1.37 [1.19–1.59] | Moderate | Strong | Strong | Moderate |
| Liew 2016a | ASD | 1.19 [1.04–1.35] | Moderate | Strong | Strong | Moderate |
| Liew 2016b | ADHD | 1.50 [1.00–2.50] | Moderate | Strong | Strong | Moderate |
| Liew 2019 | ADHD | 1.34 [1.05–1.72] | Moderate | Moderate | Strong | Moderate |
| Luo 2026a | ADHD | 1.23 [1.20–1.27] | Moderate | Strong | Moderate | Moderate |
| Luo 2026a | ASD | 1.17 [1.13–1.20] | Moderate | Strong | Moderate | Moderate |
| Luo 2026b | ADHD | 1.01 [0.93–1.08] | Strong | Strong | Moderate | Moderate |
| Luo 2026b | ASD | 1.00 [0.91–1.11] | Strong | Strong | Moderate | Moderate |
| Okubo 2025a | ADHD | 1.22 [1.06–1.41] | Moderate | Strong | Moderate | Moderate |
| Okubo 2025a | ASD | 1.06 [0.98–1.15] | Moderate | Strong | Moderate | Moderate |
| Okubo 2025b | ADHD | 0.86 [0.52–1.44] | Strong | Strong | Moderate | Moderate |
| Okubo 2025b | ASD | 0.85 [0.64–1.13] | Strong | Strong | Moderate | Moderate |
| Pleau 2026 | ADHD | 1.09 [0.94–1.27] | Moderate | Strong | Strong | Moderate |
| Prahm 2026a | ASD | 1.03 [0.95–1.12] | Moderate | Strong | Strong | Moderate |
| Prahm 2026b | ASD | 1.09 [0.91–1.27] | Strong | Strong | Strong | Strong |
| Smith Webb 2023 | ADHD | 1.80 [0.60–5.50] | Weak | Moderate | Strong | Weak |
| Stergiakouli 2016 | ADHD | 1.31 [1.16–1.49] | Moderate | Strong | Strong | Moderate |
| Sznajder 2022 | ADHD | 1.21 [1.01–1.45] | Moderate | Moderate | Moderate | Moderate |
| Tovo 2018 | ADHD | 1.42 [1.06–1.92] | Moderate | Strong | Strong | Moderate |
| Ystrom 2017 | ADHD | 1.12 [1.02–1.24] | Moderate | Moderate | Strong | Moderate |

*Thirty-nine estimates from 24 articles are reported below (family-design articles may contribute a population [a] and an author-labelled sibling [b] estimate). B1 is estimate-level and design-gated. Effect ratios are reported as published (HR/RR/OR).*

**Table S4 | Stratified pooled estimates by control level (locked 16-construct mapping; random-effects REML models with Knapp–Hartung inference). Here, n denotes effect estimates.**

| Bias domain | Strong | Moderate | Weak | Interaction p |
|---|---|---|---|---|
| B1: Familial/genetic (design-gated) | 0.99 [0.97–1.00] (n=10) | 1.15 [1.10–1.20] (n=22) | 2.08 [1.57–2.75] (n=7) | <0.0001 |
| B2: Indication / maternal health | 1.11 [1.05–1.17] (n=24) | 1.42 [1.14–1.78] (n=15) | n/a (n=0) | 0.168 |
| B3: Sociodemographic / lifestyle | 1.18 [1.07–1.30] (n=27) | 1.13 [1.03–1.24] (n=12) | n/a (n=0) | 0.752 |
| Overall (weakest-link) | 0.98 [0.95–1.02] (n=4) | 1.12 [1.07–1.17] (n=28) | 2.08 [1.57–2.75] (n=7) | <0.0001 |

**Table S5 | Sensitivity of the stratified estimates (construct granularity and rating thresholds; HR/aHR estimates; leave-one-out).**

**S5a. Current-data sensitivity across construct granularities and rating thresholds (39 estimates)**

| Sensitivity specification | # conf./cut | B1 strong | B2 strong | B3 strong |
|---|---|---|---|---|
| Coarse (Gemini-3.1) | 11 | 0.99 [0.97–1.00] | 1.11 [1.05–1.17] | 1.20 [1.09–1.34] |
| Medium: locked (primary) | 16 | 0.99 [0.97–1.00] | 1.11 [1.05–1.17] | 1.18 [1.07–1.30] |
| Fine (GPT-5.5) | 19 | 0.99 [0.97–1.00] | 1.11 [1.05–1.17] | 1.22 [1.06–1.40] |
| Looser threshold | B2 ≥2/5; B3 ≥3/8 | 0.99 [0.97–1.00] | 1.11 [1.05–1.17] | 1.17 [1.07–1.27] |
| Primary threshold | B2 ≥3/5; B3 ≥4/8 | 0.99 [0.97–1.00] | 1.11 [1.05–1.17] | 1.18 [1.07–1.30] |
| Stricter threshold | B2 ≥4/5; B3 ≥5/8 | 0.99 [0.97–1.00] | 1.08 [1.03–1.13] | 1.22 [1.06–1.40] |

**S5b. HR/aHR estimates only (updated primary mapping, n = 24)**

| Domain | Strong | Moderate | Interaction p |
|---|---|---|---|
| B1: Familial/genetic | 0.99 [0.97–1.00] (n=10) | 1.13 [1.07–1.18] (n=14) | 0.0004 |
| B2: Indication | 1.09 [1.03–1.15] (n=19) | 1.05 [0.97–1.14] (n=5) | 0.516 |
| B3: Sociodemographic/lifestyle | 1.06 [1.01–1.12] (n=16) | 1.09 [0.99–1.20] (n=8) | 0.539 |

The HR/aHR restriction contained 24 estimates and yielded an overall pooled estimate of 1.08 (95% CI 1.03–1.12). B1 pooled estimates were 0.99 (0.97–1.00; n = 10) under strong control and 1.13 (1.07–1.18; n = 14) under moderate control; no HR/aHR estimates were rated weak for B1 (interaction p = 0.0004). The leave-one-out total pooled estimates ranged from 1.13 to 1.15. Complete sensitivity results are provided in Source Data (available in the Github repository).

**Figure S3 | Random-effects meta-analysis stratified by B2 (indication/maternal health) control (REML with Knapp–Hartung inference).**

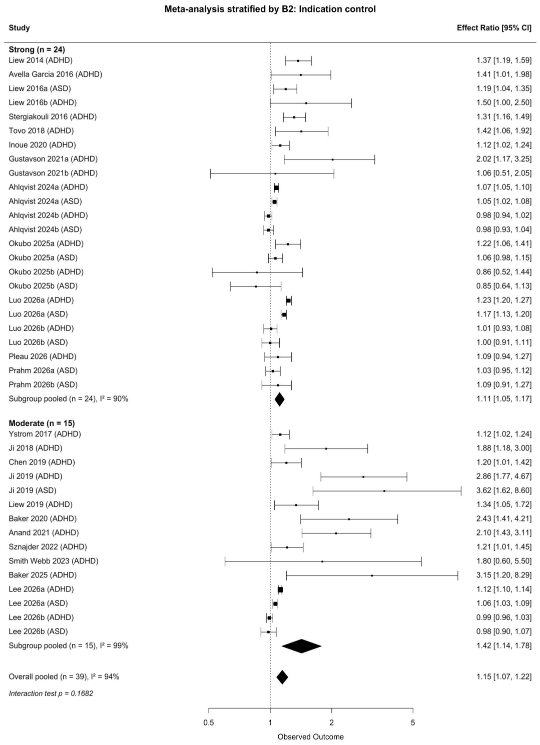


**Figure S4 | Random-effects meta-analysis stratified by B3 (sociodemographic/lifestyle) control (REML with Knapp–Hartung inference).**

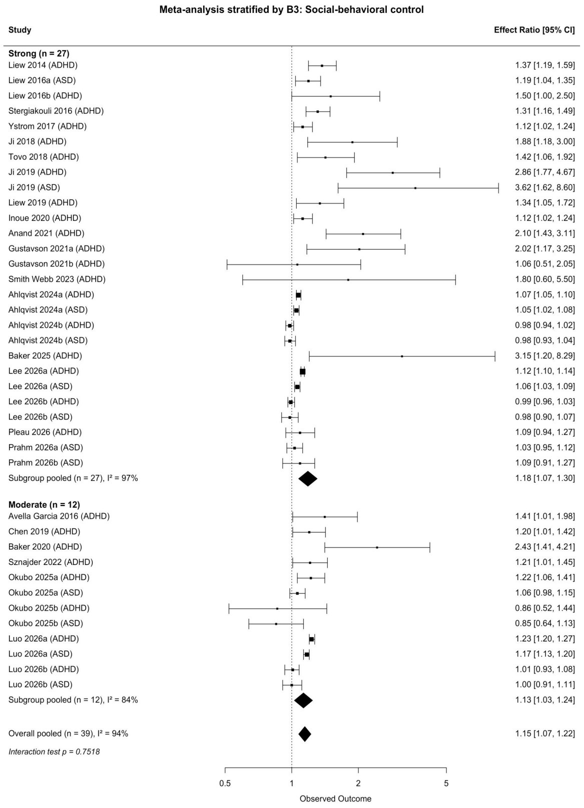

**Figure S5 | Random-effects meta-analysis stratified by overall (weakest-link) confounding control (REML with Knapp–Hartung inference).**

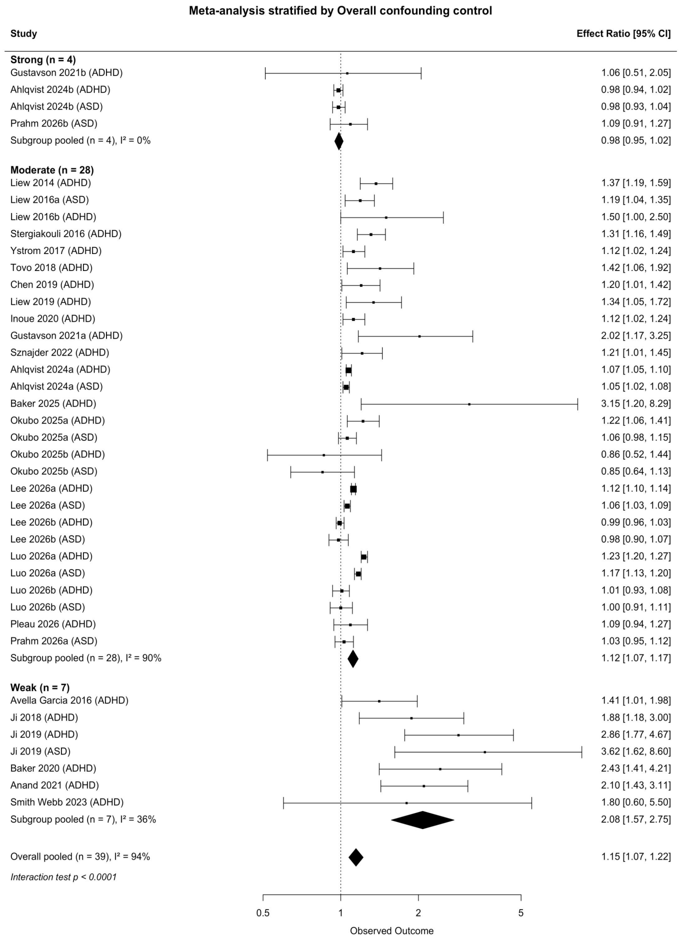

#### 5. Track B large-language-model prompts

Track B (independent construct induction) used three large language models (gpt-5.5, gemini-3.1-pro-preview, and deepseek-v4-pro), each receiving a byte-identical prompt (verified). The procedure had two phases: (1) outcome-specific candidate-variable causal-role induction of candidate confounders / mediators / colliders (three prompting strategies; outputs pooled and de-duplicated to 1,357 candidate variables); and (2) blind clustering of the pooled variables into a construct taxonomy, performed independently by each model and then reconciled against the Track A empirical audit. The models were blinded to the Track A construct scheme (no B1/B2/B3 labels and no named confounder taxonomy were shown). The two prompt specifications are reproduced below. The byte-exact prompt as sent (including all 1,357 pooled variables) and the three models' raw taxonomy outputs are provided as Source Data; API keys were read from a local environment file and appear in no distributed file.

##### Prompt 1: Outcome-specific candidate-variable causal-role induction (three prompt variants)

##### Shared block (identical in every variant, only the prompting strategy differs)

```
You are an expert in epidemiology and causal inference.

Input:
Exposure: taking paracetamol (acetaminophen) during pregnancy
Outcome: autism spectrum disorder (ASD) diagnosed in the child during childhood
Research question: the effect of taking paracetamol during pregnancy on the risk of
autism spectrum disorder in the offspring.

Your task:
Generate an exhaustive candidate list of third-party variables that may be relevant to
the causal relationship between the exposure and the outcome. Do not provide only the most obvious
variables — think broadly and systematically across all relevant epidemiologic domains.

For each variable, first locate it in time, then assign exactly one causal role.

Temporal position (relative to the exposure = paracetamol use during pregnancy, and the
outcome = ASD diagnosed in childhood):
- pre_exposure: can occur before paracetamol use begins.
- post_exposure_pre_outcome: can only occur after paracetamol use but before ASD diagnosis
  (e.g., gestational age, preterm birth, birth weight, delivery mode, breastfeeding).
- post_outcome: follows the ASD diagnosis.

Causal roles:
- confounder: a common cause of BOTH the exposure and the outcome. It is present before the
  exposure and is not caused by the exposure (timing = pre_exposure).
- mediator: a variable on the causal pathway from exposure to outcome. It is caused by the
  exposure and in turn affects the outcome (timing = post_exposure_pre_outcome).
- collider: a variable caused by both the exposure and the outcome, or a selection /
  ascertainment mechanism influenced by both.

Critical rule: a variable that can only arise after the exposure CANNOT be a confounder, even if
prior studies adjusted for it. Use the temporal position to discipline the role assignment.

Rules:
- Do not include the exposure itself or the outcome itself.
- Each variable must have exactly one role.
- Prefer specific and measurable variables.

Output only valid JSON in the following format (no prose, no markdown):
```

```
{
  "variables": [
    {
      "name": "variable name",
      "timing": "pre_exposure | post_exposure_pre_outcome | post_outcome",
      "role": "confounder | mediator | collider",
      "domain": "short category, e.g. familial/genetic, indication, socioeconomic, lifestyle, perinatal, healthcare access, environmental"
    }
  ]
}
```

**Variant A, Zero-shot**

Use the shared block exactly as above. No prompting strategy added.

**Variant B, Generic chain-of-thought**

Append to the shared block, immediately before the "Output only valid JSON" line:

```
Reason through this step by step internally before answering. Do not include your reasoning in
the response — output only the final JSON.
```

**Variant C, Framework chain-of-thought**

Append to the shared block, immediately before the "Output only valid JSON" line:

```
Framework for causal reasoning — reason internally using these steps; do NOT output the steps:
Step 1. List variables that can directly influence whether the exposure occurs (e.g., indications
        for the medication, maternal health conditions, healthcare access, prescribing behavior,
        sociodemographic and behavioral factors).
Step 2. List variables that can be directly affected by the exposure (biological, physiological,
        pregnancy-related, perinatal, or healthcare consequences occurring after the exposure).
Step 3. List variables that can directly influence whether the outcome occurs (genetic, familial,
        prenatal, perinatal, postnatal, environmental, healthcare, diagnostic, child-level factors).
Step 4. List variables that can be directly affected by the outcome (diagnostic pathways,
        healthcare use, developmental assessment, parental behavior, selection into records).
Step 5. Assign roles:
        - cause of both exposure and outcome  -> confounder
        - consequence of exposure AND cause of outcome -> mediator
        - consequence of both, or a selection/ascertainment mechanism -> collider
```

**Prompt 2: Blind construct clustering of the pooled variables**

**Track B, blind construct induction prompt**

Purpose: induce Track B's OWN construct taxonomy from the pooled LLM-generated variables,

**independently** of the empirical 17-construct pre-adjudication Track A scheme. This is the missing Stage-1B step

("Pool and harmonize candidate nodes" → "Background causal candidate graph") in Fig. 1.

Independence is the whole point: the model must NOT be shown, and must NOT reproduce, any

pre-existing classification (no B1/B2/B3, no named confounder taxonomy). The taxonomy must emerge

purely from the variables themselves. The resulting Track-B constructs are later mapped

against Track A, convergence = validation, divergence = blind spots / over-adjustment.

Input: trackB_pooled_variables.json, 1,357 unique variables, each with name, role (confounder/mediator/collider), timing (pre_exposure / post_exposure_pre_outcome / post_outcome).

DeepSeek-V4-pro used temperature 0 for both phases. GPT-5.5 used reasoning_effort=medium without an explicit temperature parameter. Gemini-3.1-pro-preview used temperature 0 for candidate-variable induction and no explicit temperature parameter for blind construct clustering.

**PROMPT**

```
You are a causal epidemiologist building a taxonomy from scratch.

Background causal question (context only, do not answer it):
  Exposure  = taking paracetamol (acetaminophen) during pregnancy
  Outcome   = autism spectrum disorder (ASD) diagnosed in the child during childhood

You are given a long list of candidate third-party variables that multiple experts independently
proposed for this exposure-outcome question. Each variable carries a causal role (confounder /
mediator / collider) and a temporal position relative to the exposure and the outcome.

YOUR TASK
Inductively group these variables into a parsimonious set of higher-level CONSTRUCTS, purely from
the variables themselves.

A CONSTRUCT is a single latent causal factor that several specific measured variables are proxies
or instances of. Example pattern (illustrative, not from this list): many specific infections,
fevers, and inflammatory markers all instantiate one underlying "acute maternal infection"
construct. A construct is broader than a single measured variable, but narrower than a whole
thematic domain.

INDEPENDENCE, STRICT
- Build the taxonomy ONLY from the variables given.
- Do NOT use, name, or map onto any pre-existing or standard classification scheme, framework,
  or domain labels. Let the structure emerge from the data alone.
- Do not invent variables that are not in the list.

GRANULARITY
- Aim for mid-level constructs: each a distinct causal mechanism or common cause.
- As a rough guide expect on the order of 15-35 constructs, but choose whatever number best fits;
  merge near-synonyms aggressively, split only when two groups are genuinely causally distinct.
- Cluster ALL variables, regardless of role. Mediators, colliders, and selection/ascertainment
  variables must form their OWN constructs too (do not drop them).

FOR EACH CONSTRUCT, report:
- a short descriptive name (your own wording);
- a one-sentence causal definition (what underlying factor it represents);
- dominant_role: the causal role most of its members share (confounder / mediator / collider /
  mixed), inferred from the members;
- typical_timing: pre_exposure / post_exposure_pre_outcome / post_outcome / mixed;
- member_variables: the exact names of the input variables assigned to it (every input variable
  must be assigned to exactly one construct).

OPTIONAL, after building the constructs, you MAY propose higher-level groupings of your own
constructs IF a natural super-structure emerges, but do not force it and do not use any standard
domain names.
```

```
Output ONLY valid JSON (no prose, no markdown):
{
  "constructs": [
    {
      "id": "C1",
      "name": "construct name",
      "definition": "one-sentence causal definition",
      "dominant_role": "confounder | mediator | collider | mixed",
      "typical_timing": "pre_exposure | post_exposure_pre_outcome | post_outcome | mixed",
      "member_variables": ["exact variable name", "..."]
    }
  ],
  "optional_super_groups": [
    { "name": "group name", "construct_ids": ["C1", "C4"] }
  ]
}

INPUT VARIABLES (name | role | timing):
<<< the 1,357 pooled variables are inserted here, one per line >>>
```

**Notes on running**

- The model must assign all 1,357 variables; if output truncates (finish_reason=length), split the input into 2 halves with the SAME instruction, induce a taxonomy on each, then reconcile, or switch to a taxonomy-only pass (constructs + 5 example members each) and assign the full 1,357 in a cheap second classification pass against the induced taxonomy.

- Run the identical prompt on gpt-5.5, deepseek-v4-pro, and gemini-3.1-pro-preview→ three independently-induced Track-B taxonomies. Their mutual agreement is itself a robustness check before mapping to Track A.

- freq (cross-run consensus count) is deliberately withheld from the model so clustering is driven by causal meaning, not popularity; we keep freq for later weighting.

#### 6. A reusable protocol for applying the framework to other exposure–outcome questions

The procedure used here is not specific to prenatal paracetamol. This section restates it as a transferable, step-by-step protocol, so that the same bias-domain triangulation can be applied to other exposure–outcome questions in which observational estimates fail to converge. Placeholders written as {EXPOSURE} and {OUTCOME} are replaced with the target question. The prompts themselves are not reprinted here: the prompts used in this study are given in Section 5, and the protocol indicates where the exposure and outcome are substituted. Supplementary Table S6 summarises the six stages.

**Table S6.** Summary of the bias-domain triangulation protocol.

| Stage | Purpose | Process |
| --- | --- | --- |
| **1. Track A audit** | Record what the focal literature actually controlled for, as constructs. | 1. Select each study's main adjusted model, the one pooled.<br>2. Extract its covariates; exclude sensitivity-, stratified-, mediation- or selection-only variables.<br>3. Harmonise raw variables into constructs.<br>4. Double-extract and adjudicate. |
| **2. Track B background DAG** | Derive what should be considered, independently of Track A. | 1. Query at least three LLMs from independent developers, using the model-specific generation parameters reported in Section 5, blind to Track A.<br>2. Three strategies: zero-shot, generic and framework chain-of-thought.<br>3. Each variable gets one causal role and a temporal position.<br>4. Pool, de-duplicate, and induce a construct taxonomy. |
| **3. Integration** | Reconcile the two tracks into one locked construct set. | 1. Adopt the fullest Track B taxonomy as the spine.<br>2. Cross-walk to Track A: convergent, blind-spot, or role-conflicting.<br>3. Adjudicate roles by admissibility criteria.<br>4. Separate core (scored) from non-core. |
| **4. Bias domains** | Group core confounders into a few domains. | 1. Derive the domains per problem from the core confounders.<br>2. Separate the design-controlled latent domain from the measurement-controlled domains; split the measured set by mechanism.<br>3. Pre-register the scheme; the number and partition of domains are modelling choices, tested for robustness. |
| **5. Scoring** | Rate each estimate's control per domain. | 1. Count-based domains: Strong / Moderate / Weak by a pre-registered threshold.<br>2. Latent domain: a design-gated rule.<br>3. Aggregate by the weakest-link rule.<br>4. Test sensitivity to thresholds and granularity. |
| **6. Read-out** | Read attenuation as a causal-structure signal. | 1. Pool with random-effects REML models using Knapp–Hartung inference. / 2. Examine the within-domain gradient toward the null. / 3. Compare strong-control estimates across domains. / 4. Restrict by effect-measure family and run leave-one-out diagnostics. |

**Scope.** The protocol applies when the evidence is observational and reports ratio-type estimates poolable on the log scale; when repeated syntheses have not converged; when the exposure is plausibly structured by background liability, clinical indication, health-seeking behaviour, or social context; and when the study pool is heterogeneous in adjustment and ideally contains a few design-based studies (sibling, family fixed effects, twin, or negative-control) to anchor a latent-liability domain. The pipeline has six stages; Stages 1 and 2 run in parallel and stay blind to each other.

**Stage 1. Track A, empirical adjustment-set audit.** For each study, extract covariates from the single model whose estimate enters the meta-analysis, the authors' primary fully adjusted model, excluding variables used only in sensitivity, stratified, mediation, or selection analyses. For sibling or family-design papers reporting both a population-adjusted and a within-family estimate, record both as separate rows. Harmonise raw variables into constructs, so that a construct counts as controlled whenever any adequate proxy for it is adjusted. Two annotators extract independently; quantify agreement and resolve discrepancies against the source PDFs.

**Stage 2. Track B, independent background DAG. Independently of Track A, reverse-engineer the background causal structure with at least three large language models from independent developers, none of which sees the Track A constructs or the bias-domain scheme. The exact prompts are those in Section 5: Prompt 1 (outcome-specific candidate-variable causal-role induction under the three reasoning scaffolds, zero-shot, generic chain-of-thought, and framework chain-of-thought, using the model-specific generation parameters reported in Section 5) and Prompt 2 (blind construct induction over the pooled variables). To apply the framework to a different question, substitute {EXPOSURE} and {OUTCOME} for the exposure and outcome wherever they appear in those prompts, and keep the temporal-discipline and independence instructions unchanged. Pool the returned variables, de-duplicate them, and have each model induce its own construct taxonomy from the pooled list.**

**Stage 3. Integration and causal-role adjudication.** Adopt the most complete Track B taxonomy as the spine after verifying it subsumes the others, and cross-walk it onto Track A, labelling each construct convergent (both sources), blind-spot (Track B only), or role-conflicting. Adjudicate each construct's causal role against pre-specified admissibility criteria (temporality, biological or causal plausibility, back-door relevance, adjustment admissibility, and cross-source concordance), and separate core pre-exposure common causes, which are scored, from non-core constructs (mediators, colliders, instruments, and precision covariates), which remain in the matrix for transparency but are not scored. Track-B-only constructs that the focal literature rarely measures are reported as blind spots but not scored.

**Stage 4. Defining bias domains.** Group the core confounders into bias domains. Neither the number of domains nor the way constructs are partitioned among them is unique: it is a pre-registered modelling choice that trades attribution against power. A finer scheme localises bias to a more specific axis but spreads the estimates thinly across strata; a coarser scheme is better powered but attributes bias more bluntly. A handful of domains, often two to four, is usually a workable balance, and the choice should be tested for robustness by re-running under coarser and finer schemes (Stage 5). What generalises is the organising principle, not a fixed list or a fixed count: two constructs belong in the same domain when they share both a bias-generating mechanism and the means required to close it, design or measurement. The one invariant is to separate the latent or structural common causes, which can be addressed only by design (a sibling or fixed-effects comparison, a negative control, and so on), from the measured common causes that regression can adjust; within the measured set, the partition into mechanism-based domains (commonly drivers of exposure assignment and background context) is itself one reasonable scheme among several. The content of each domain is problem-specific. In particular, the latent domain is shared familial and genetic liability for a heritable outcome such as the one studied here, but it may instead be a shared environment, a clinic or site, a region, or a secular time trend in other questions, where familial and genetic liability need not appear at all. Supplementary Table S7 gives common archetypes and the rating rule; the three-domain instantiation used in this study (B1 familial/genetic liability, B2 clinical indication and maternal health, B3 social-behavioural context) is given in main-text Table 1.

**Stage 5. Scoring confounding control.** Rate each estimate Strong, Moderate, or Weak within every domain, then aggregate. The generic rating rule is given in Supplementary Table S7: a count-based rule for measurement-controllable domains (Strong when a substantial share of the domain's constructs is controlled) and a design-gated rule for a latent domain (Strong only for an eligible design such as sibling or family fixed effects). A construct counts as controlled when any adequate proxy for it is adjusted. Aggregate the domain ratings with a conservative weakest-link rule: Strong only if every domain is Strong, Weak if any domain is Weak, Moderate otherwise. The exact thresholds applied in this study (for example three of five and four of eight constructs) are given in main-text Table 1 and the Online Methods. Because thresholds and construct granularity are modelling choices, repeat the rating under stricter and looser thresholds and coarser and finer granularities, and report which conclusions are invariant.

**Table S7.** Generic bias-domain template and rating rule, organised by the control mechanism that generalises across questions.

| Domain archetype | What it captures (content is problem-specific) | Strong | Moderate | Weak |
|---|---|---|---|---|
| Latent / structural common cause (design-controlled) | Unmeasured shared causes regression cannot reach, e.g. familial or genetic liability (heritable outcomes), shared environment or household, clinic, site or region, or secular time trend. | An eligible design that differences out the latent cause (sibling or twin comparison, family or unit fixed effects, negative control) | A population model adjusting at least one measured proxy of the latent factor | Neither a design-based control nor a measured proxy |
| Drivers of exposure assignment (measurement-controlled) | Why the exposure occurs, e.g. clinical indication, channeling, provider behaviour, selection into exposure, reverse-causation drivers. | A substantial share of the domain's constructs controlled (pre-registered cut) | At least one construct controlled, but below the Strong cut | No construct in the domain controlled |
| Background context (measurement-controlled) | Broad observed background, e.g. socioeconomic position, demographics, lifestyle, psychosocial factors. | A substantial share of the domain's constructs controlled (pre-registered cut) | At least one construct controlled, but below the Strong cut | No construct in the domain controlled |

**Stage 6. Stratified meta-analysis and triangulation read-out.** Pool with random-effects models (REML, Knapp–Hartung), combining hazard, risk and odds ratios on the log scale, and test subgroup differences with a categorical-moderator Q-test. Read three things in order: the unstratified pool, confirmed by leave-one-out; the within-domain gradient, asking whether each domain's pooled estimate moves toward the null as control strengthens; and the cross-domain comparison at the strong tier. If strong control of one domain reaches the null while the others do not, the evidence is more compatible with that domain's bias structure than with a population-level effect. If all domains converge to the null as control strengthens, the observed association is more compatible with being largely explained by confounding, although shared residual bias and statistical uncertainty remain possible. Attenuation that tracks one bias axis specifically is the triangulation signature.
